# Revisiting acyclovir dosing for viral encephalitis using a Bayesian PBPK modeling approach

**DOI:** 10.1101/2024.08.25.24312421

**Authors:** Ming Sun, Martijn L. Manson, Anne-Grete Märtson, Jacob Bodilsen, Elizabeth C.M. de Lange, Tingjie Guo

## Abstract

Acyclovir is a primary treatment for central nervous system (CNS) infections caused by herpes simplex virus (HSV) and varicella-zoster virus (VZV). However, patient outcomes remain suboptimal with high mortality and morbidity, following current dosing guidelines. Given the lack of alternative therapies, there is a pressing need to optimize acyclovir dosing, especially since initial regimens were developed in the 1980s with incomplete pharmacokinetic data in the CNS. This study aimed to evaluate both current and alternative acyclovir dosing regimens using a full Bayesian physiologically-based pharmacokinetic (PBPK) model tailored for viral encephalitis. We developed a CNS PBPK model to simulate acyclovir concentrations in plasma, brain extracellular fluid (ECF), and subarachnoid space (SAS). Drug efficacy was assessed using two pharmacokinetic targets, 50%*f*T>IC_50_ and C_min_>IC_50_, with a safety threshold set at 25 mg/L in plasma. The standard dosing regimen (10 mg/kg TID) yielded sufficient acyclovir exposure in plasma, brain extracellular fluid (ECF), and subarachnoid space (SAS) compartments based on the 50%*f*T>IC_50_ target. However, it did not consistently meet the C_min_>IC_50_ target, indicating potential suboptimal exposure in these compartments when evaluated against this criterion. Notably, a higher probability of target attainment (PTA) was generally observed in the brain ECF and SAS compared to plasma. Increasing the dosing frequency to QID improved target attainment but exceeded the toxicity threshold at 20 mg/kg. Our findings suggest that a dosing regimen of 10 mg/kg or 15 mg/kg QID may offer a more effective and safer approach for managing CNS infections compared to the other tested alternative dosing regimens.

## Introduction

Viral encephalitis is a severe inflammation of the central nervous system (CNS) caused by viral infections, often involving herpes simplex virus (HSV) and varicella-zoster virus (VZV). The disease is rare but fatal and survivors often encounter severe neurological impairment despite drug treatment [1]. Acyclovir is the primary drug used in clinical practice to treat these infections, with a standard dosing regimen of 10 mg/kg administered intravenously three times daily (TID) for HSV encephalitis, and up to 15 mg/kg for VZV encephalitis [2]. Although standard dosing of acyclovir significantly reduces mortality compared to untreated cases, the mortality and morbidity for both HSV encephalitis and VZV encephalitis patients remain high [3]. Clinical guidelines recommend early initiation of acyclovir treatment upon hospital admission to enhance treatment outcomes [4-6]. However, substantial mortality and morbidity have also been observed in patients receiving early acyclovir therapy [7, 8], suggesting that the current acyclovir dosing might need to be revisited. The current clinical dosing of acyclovir for HSV encephalitis was established based on limited evidence, primarily from two clinical trials conducted in the 1980s [9, 10], while for VZV encephalitis there has been no clinical trials conducted so far. In these studies, the chosen dosing regimen of 10 mg/kg TID was based on the plasma pharmacokinetics (PK) of acyclovir observed after a single intravenous infusion [11]. However, in contrast to the systemic exposure of acyclovir, there has been limited investigations into the PK of acyclovir’s within the CNS. Therefore, it is unclear whether drug concentrations in plasma are representative for the exposure of acyclovir within the CNS, the target site of action in treating viral encephalitis. This knowledge gap raises concerns about whether the current dosing regimen effectively ensures sufficient CNS exposure to achieve optimal therapeutic outcomes for viral encephalitis.

Given the uncertainties surrounding acyclovir’s CNS exposure, there is a critical need to reassess and optimize dosing regimens specifically for CNS viral infections. Several model-based approaches using PK targets such as maintaining acyclovir concentrations above the IC_50_ for 50% of the dosing interval (50%*f*T>IC_50_) or ensuring the trough concentration (C_min_) exceeds the IC_50_ (C_min_>IC_50_), have been developed for systemic viral infections [12-14]. However, these methods, which rely heavily on drug exposure data to facilitate model construction, may not be entirely applicable for describe the complexities of drug CNS PK due to the limited information on acyclovir CNS exposure. This limitation highlights the need for reassessment and optimization of acyclovir dosing regimens specifically for CNS viral infections, potentially through alternative modeling approaches that better account for the unique PK properties of the CNS.

The use of physiologically-based pharmacokinetic (PBPK) modeling, particularly when combined with Bayesian methods, offers a promising approach to better understand drug distribution within the CNS and address the limitations posed by sparse data [15]. Compared to the empirical PK modeling approaches, PBPK models differentiate system-specific and drug-specific parameters to allow for scaling and translation of the model prediction across species (such as from preclinical to clinical), and disease conditions. Additionally, due to a knowledge-driven nature of model building procedure, PBPK modeling has the advantage of analyzing scarce PK data while possessing a complete predictive capability. Due to highly restriction on obtaining the samples from the brain, PK data from human brain is sparsely available and PBPK modeling can therefore be a suitable alternative approach to predict PK within different CNS regions. PBPK modeling allows for the mechanistic description of the physiology and therefore can provide an informative insight into the drug distribution in the CNS. In addition, the Bayesian method has been lately proposed as an addition to the PBPK modeling to allow for integration of existing knowledge into the analysis to further optimize the predictive capability of the model. In a Bayesian analysis, prior knowledge, e.g., from existing literature, can be included as an extra source of data for the model fitting to allow for the characterization of the parameters’ uncertainty, which allows us to better understand the real-world variability.

To address the gap in understanding acyclovir CNS pharmacokinetics, we set out to study acyclovir CNS PK using a full Bayesian PBPK approach. Facilitated by the developed model, we also aimed to evaluate the current and alternative acyclovir dosing regimens for the optimal treatment of CNS viral infection.

## Methods

In this study, we performed the modeling based on an existing CNS PBPK model framework (LeiCNS 3.0) [16] using published literature data of acyclovir following a full Bayesian approach. The developed model was used to simulate the CNS PK profile of the current standard acyclovir dosing regimen as well as three alternative dosing regimens. Based on the predefined therapeutic target and toxicity threshold, we evaluated the suitability of these acyclovir dosing regimens for treating viral encephalitis (Figure 1).

**Figure 1.**
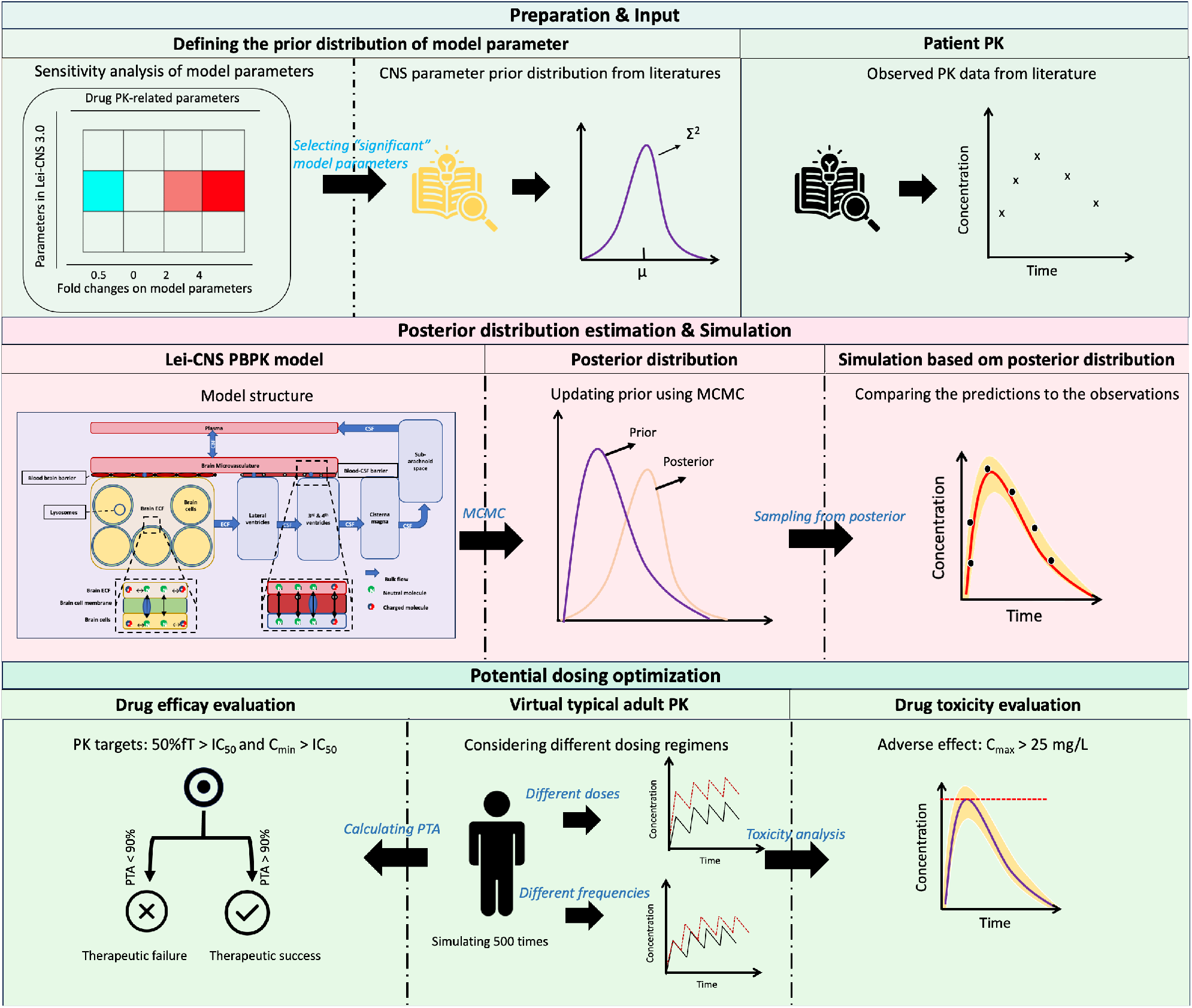
Overview of the approaches. μ and ∑^2^, the mean and the variance of the selected parameter; 50%*f*T > IC_50_, drug concentrations at the steady state >IC_50_ for 12h during 24 h treatment; C_min_ > IC_50_, drug trough concentration at the steady state during the whole treatment; MCMC, Markov chain Monte Carlo; PTA, probability of drug target attainment.

### PK data and LeiCNS 3.0 model framework

Our modeling framework was based on a previously developed CNS PBPK model (LeiCNSPK3.0), which characterizes PK of a series of drugs in nine CNS compartments including brain microvessels, brain extracellular fluid (ECF), brain intracellular fluid (ICF), lysosomes, lateral ventricles, third and fourth ventricles, cisterna magna, and subarachnoid space (SAS) [16]. The model consisted of parameters of drug-specific properties and system-specific properties collected from various sources of data such as published literature data, or in-house microdialysis studies (Table S1). For the analysis, we used acyclovir concentration data from both plasma and the lumbar site (subarachnoid space, SAS), identified from two studies [17, 18].

### Bayesian analysis

The general workflow involves three key steps: 1) sensitivity analysis, 2) defining the prior distributions of model parameters, and 3) estimating the posterior distribution of parameters (Figure 1).

#### 1) Identifying sensitive parameters

A sensitivity analysis (SA) was first performed to identify systemic parameters (excluding drug-specific ones) to which the model is sensitive. A 0.1- to 10-fold change was applied to all system-specific parameters and the corresponding change in the model-predicted drug concentrations was examined. The selection of model parameters for further Bayesian analysis was based on whether the impact of the parameter in question on the predicted area under the concentration-time curve (AUC) exceeded 15% in any model compartment. The identified sensitive parameters were set free for estimation in the Bayesian analysis and the remaining parameters were fixed at the reported value (Table S1).

#### 2) Defining prior distribution

For the identified sensitive parameters, we set an prior using log-normal distributions according to literature data (Table S2). Likelihood functions for plasma and subarachnoid space observations were modeled assuming a log-normal distribution and the prior for the variance of the residuals were set using a half-Cauchy distribution.

#### 3) Estimating posterior distribution

The Markov chain Monte Carlo (MCMC) method with the No-U-turn sampler (NUTS) was used to estimate the posterior distributions of model parameters based on an acceptance rate of 0.8. Four Markov chains were deployed in parallel with 1000 iterations each, including 500 iterations of warmup. The Gelman-Rubin statistic R^ and normalized effective sample size (Neff/N) were calculated to evaluate the convergence and efficiency of MCMC sampling process, respectively. Additionally, MCMC sampling trace plots and posterior distributions were used to visually examine the convergence and robustness of the analysis. Finally, posterior predictive checks (PPC) were consulted to assess the predictive performance of the model.

### Dose regimens evaluation

#### Efficacy

Effective concentrations of acyclovir for treating CNS viral infections were defined using two PK targets: 1) maintaining acyclovir concentrations above the half-maximal inhibitory concentration (IC_50_) for at least 50% of the time, denoted as 50%*f*T>IC_50_ [12, 13], and 2) ensuring trough concentrations (C_min_) to exceed the IC_50_, [14]. For this study, IC_50_ values were set at 0.56 mg/L for HSV and 1.125 mg/L for VZV [13].

To evaluate the adequacy of different dosing regimens in achieving these effective concentrations, we conducted a probability of target attainment (PTA) analysis. The PTA was calculated as the percentage of simulations where the defined PK targets were achieved (Equation 1).

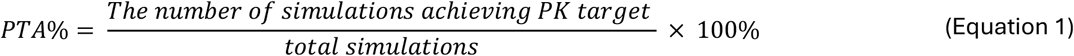

We considered two distinct PK targets: 50%*f*T>IC_50_ and C_min_>IC_50_, where a 90% threshold was set to ensure a high likelihood of achieving therapeutic efficacy while minimizing the risk of resistance development. Using the developed model, we simulated acyclovir concentrations in plasma, brain ECF, and SAS based on 500 posterior samples i.e. resulting in 500 times simulation. A range of dose regimens were considered in the simulation including the current standard dosing regimen (10 mg/kg/8h) and alternative regimens of 10 mg/kg, 15 mg/kg, and 20 mg/kg administered either three times daily (TID) or four times daily (QID).

#### Toxicity

Previous study has identified a plasma toxicity threshold of 25 mg/L for acyclovir, above which there is an increased risk of renal dysfunction and neurotoxicity [19]. As such, we evaluated predicted acyclovir concentrations in plasma for 15 mg/kg and 20 mg/kg (TID or QID), against a toxicity threshold of 25 mg/L. Plasma concentrations surpassing this threshold were considered to represent a higher risk of toxicity.

### Software and data analysis

The acyclovir concentration data were extracted using WebPlotDigitizer (version 4.2). The PBPK modeling and Bayesian analysis were performed using Stan (version 2.27.0) and its R interface CmdStanR (version 0.61), facilitated by Torsten (version 0.89). Data visualization was performed using R (version 4.3.3) using R packages ggplot2 (version 3.4.4) and bayesplot (version 1.10.0).

## Results

### Model development and evaluation

We identified eight sensitive parameters which were carried forward to the Bayesian analysis (Figure S1, Table S2). We successfully estimated the posterior distribution for all the selected model parameters (Table S3). The numerical results showed that all R^ values were close to 1 indicating a successful convergence of the parameter estimations. All Neff/N values exceeded 0.5, suggesting that the model parameters were reliably estimated (Figure S2). Trace plots showed robust mixing of posterior samples indicated by their “fuzzy caterpillar” appearance. The overlaid density plots from all chains demonstrated convergence to a consistent distribution (Figure S3A). Comparing to the prior, the posterior distribution of parameters became more condensed and informative, with uncertainty around 1.5 to 2-fold from the mean values, indicating improved precision and reliability in the model parameters (Figure S3B). The posterior predictive check showed that the model was able to adequately predict the observed data, with most observations falling within the 95% prediction interval (Figure 2).

**Figure 2.**
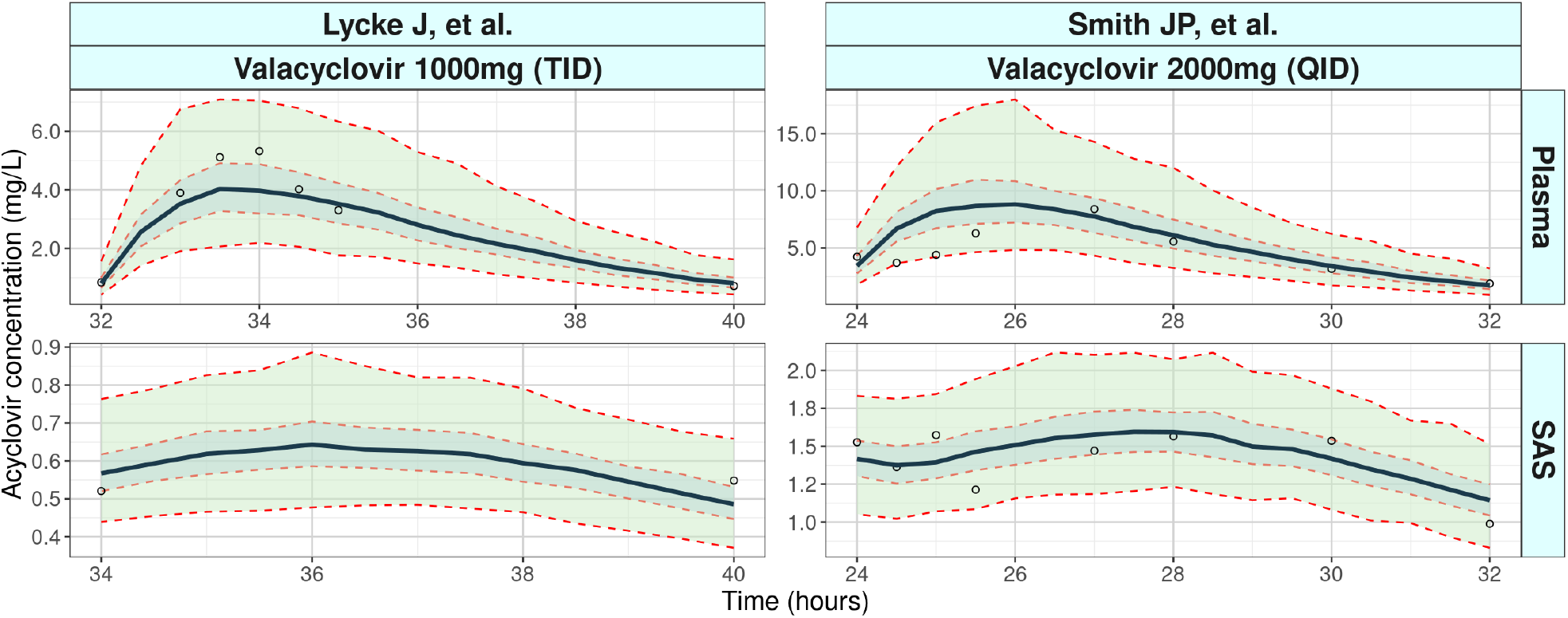
Posterior predictive check (PPC) for the Bayesian CNS PBPK model. Simulated acyclovir data over time in plasma and brain subarachnoid space (SAS) are shown (Line, median; middle band color, 50% interval; outer band, 95% interval) against the observations (Dots, mean values).

### PK profiles of acyclovir under the standard dosing regimen

The PK data of acyclovir identified in the literature were based on oral administration of valacyclovir which is a prodrug of acyclovir. We therefore corrected the valacyclovir-acyclovir conversion in the model eliminating absorption and biotransformation for any simulation-based process. We simulated acyclovir concentrations in plasma, brain ECF, and SAS under the standard IV dosing regimen (10 mg/kg, TID). The pharmacokinetic (PK) profiles (Figure 3) indicate distinct differences among these compartments.

**Figure 3.**
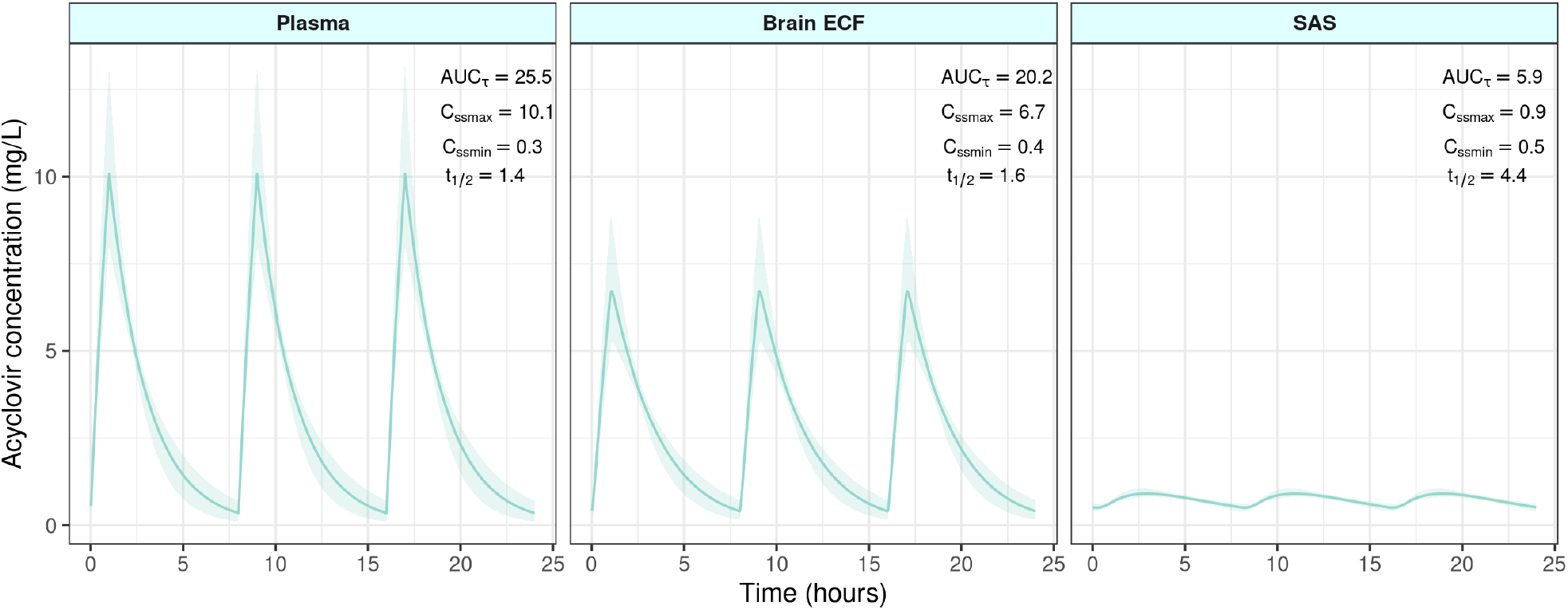
Simulated acyclovir concentrations over time in different compartments (plasma, brain extracellular fluid (ECF), and subarachnoid space (SAS)) for the standard dosing regimen of 10 mg/kg administered three times daily (TID). The solid lines represent median predicted concentrations, with the shaded areas indicating the 95% confidence intervals. The area under the curve over 8 hours (AUC_τ_), the maximum concentration at the steady state (C_ssmax_), the trough concentration (C_ssmin_), and the half-life (t_1/2_) were calculated based on the predicted concentrations in each compartment.

In the plasma compartment, the highest drug exposure was observed, with an area under the concentration-time curve (AUC) over 8 hours, which measures the total drug exposure over the dosing interval, reaching 25.5 mg·h/L. The peak concentration (C_ss,max_) was 10.1 mg/L, and the drug was rapidly eliminated, as indicated by a half-life (t_1/2_) of 1.4 hours. In contrast, both CNS compartments, brain ECF and SAS, showed lower AUC and C_ss,max_ but higher trough concentrations (C_ss,min_) (Figure 3). The SAS compartment exhibited the lowest drug exposure with an AUC of 5.9 mg·h/L and a C_ss,max_ of 0.9 mg/L, while having the highest C_ss,min_ of 0.5 mg/L and a much longer half-life of 4.4 hours.

### PTA of acyclovir based on plasma and brain ECF concentrations

#### Efficacy

The efficacy of acyclovir was first assessed using the PK target of 50%*f*T>IC_50_ (Figure 4A). The standard dosing regimen of 10 mg/kg TID achieved a PTA exceeding 90% for HSV infection across all compartments, including plasma, brain ECF, and SAS. However, for VZV infection, this threshold was not met in the SAS compartment until the dosage was increased to 20 mg/kg. Increasing the dosing frequency to QID significantly improved PTA outcomes for the 50%*f*T>IC_50_ target. For HSV, all doses ranging from 10 to 20 mg/kg achieved a PTA exceeding 90% across all compartments. For VZV, the PTA exceeded 90% with all doses from 10 mg/kg to 20 mg/kg QID in both plasma and brain ECF. However, the 10 mg/kg QID regimen did not achieve the 90% PTA threshold in the SAS compartment.

**Figure 4.**
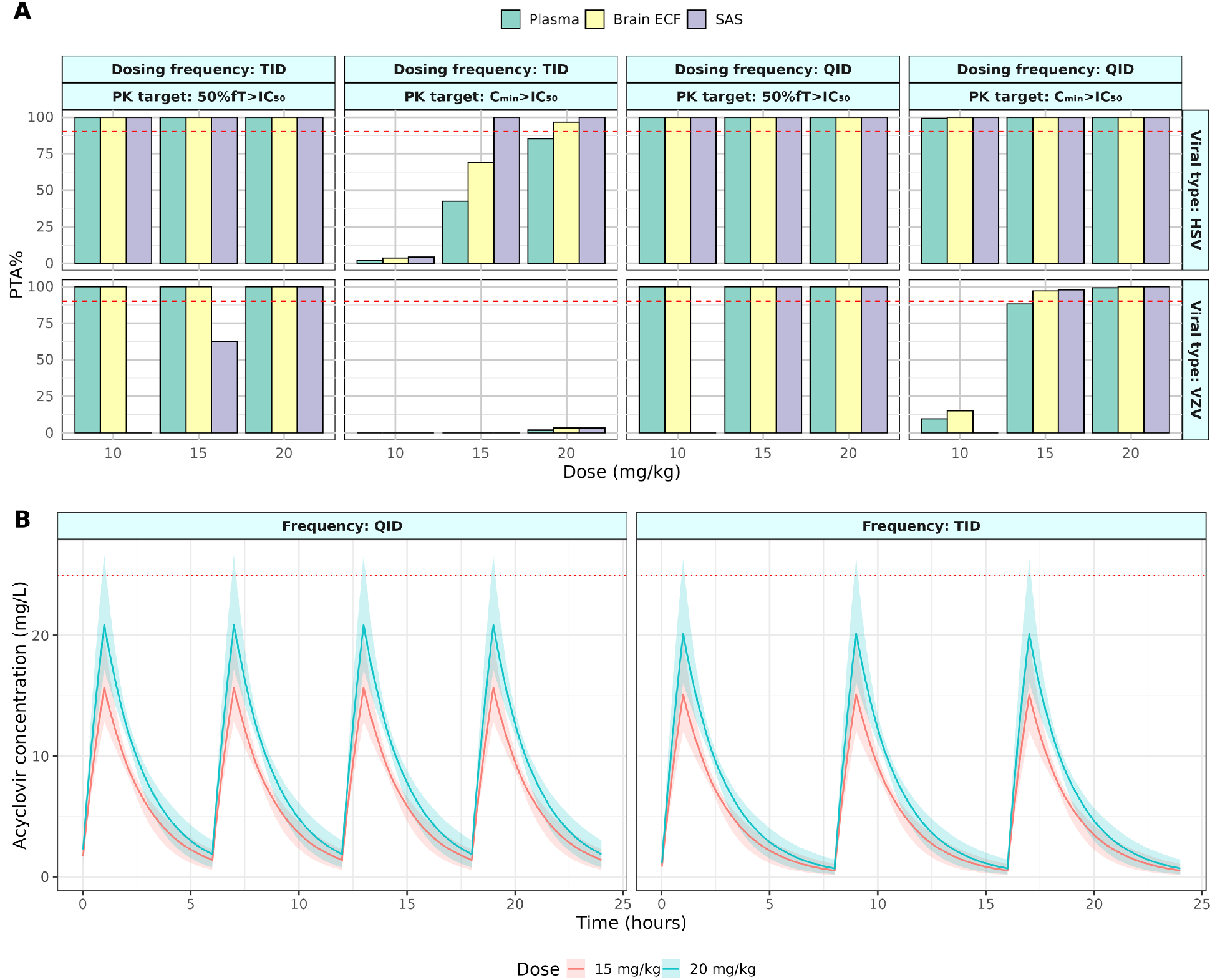
(A). Probability of target attainment (PTA) of acyclovir in treating herpes simplex virus (HSV) and varicella-zoster virus (VZV) in plasma, brain extracellular fluid (ECF), and subarachnoid space (SAS) at different doses (10 mg/kg, 15 mg/kg, 20 mg/kg) and dosing frequencies (TID: three times per day; QID: four times per day). The red dashed line marks the PTA of 90%, the threshold defined to indicate adequate acyclovir dose regimens. (B). Simulated plasma concentrations of acyclovir over time for different dose regimens (15 to 20 mg/kg with TID or QID dosing). The solid line represents the median value of predicted concentration, with the shaded area representing the 95% confidence interval. The red dashed line represents the threshold concentration (25 mg/L) associated with moderate to severe side effects.

When using the PK target of C_min_>IC_50_, the PTA for the standard 10 mg/kg TID regimen was below 5% across all compartments, indicating suboptimal treatment for both HSV and VZV (Figure 4A). For HSV, only the higher doses of 15 mg/kg and 20 mg/kg TID reached a PTA of 90% in the SAS and brain ECF compartments. For VZV, the PTA remained below 4% across all compartments, even at the 20 mg/kg TID dose. Increasing the dosing frequency to QID also enhanced PTA outcomes for the C_min_>IC_50_ target. For HSV, all doses from 10 to 20 mg/kg surpassed the 90% PTA threshold in all compartments. In the case of VZV, the 15 mg/kg and 20 mg/kg QID regimens either approached or achieved a PTA of 90% across each compartment. However, the 10 mg/kg QID dose failed to meet this target in any compartment, highlighting a continued suboptimal response in achieving adequate trough concentrations.

#### Toxicity

The results showed that a dose of 20 mg/kg, whether administered TID or QID, resulted in acyclovir plasma concentrations exceeding the toxicity threshold of 25 mg/L at peak concentration during steady state (Figure 4B), suggesting a risk of adverse effects. Conversely, doses of 15 mg/kg or below, regardless of dosing frequency (TID or QID), were considered safe, as they did not surpass the defined toxicity threshold.

## Discussion

Acyclovir has been commonly used as a first-line treatment for HSV and VZV associated viral encephalitis. However, our understanding of acyclovir exposure within the CNS and whether this exposure corresponds to the theoretically required levels is limited, suggesting a need to revisit current acyclovir dosing regimens. In this study, we performed a full Bayesian PBPK modeling for characterizing acyclovir CNS distribution for the first time which allowed us to optimize acyclovir dosing.

We found that the PTA can differ significantly between the choice of PK targets. An appropriate PK target is probably dependent on both the specific antiviral agent and the type of virus [20]. For HSV and VZV infection, the 50%*f*T>IC_50_ target and the C_min_>IC_50_ target are both commonly employed in model-informed approaches for guiding acyclovir dosing. Our results showed that with the current standard dosing regimen, the PK target 50%*f*T>IC_50_ can be achieved in general, whereas C_min_>IC_50_ cannot be consistently met. This aligns with our expectation as the C_min_>IC_50_ target is stricter by definition where drug concentration needs to be always above IC_50_ to be on target. Nonetheless, these targets are solely based on PK which miss the relationship of acyclovir exposure and its antiviral effect. As such, understanding the antiviral pharmacodynamics (PD) is a logical next step to further improve acyclovir dosing rationality. For instance, viral dynamics has been increasingly studied in antiviral treatment optimization. Model-informed dose optimization can be extended from PK modeling to PK/PD modeling to link drug concentration with viral load reduction. This will allow us to derive optimal dose regimens that are directly responsible to the antiviral effect within the brain. Currently, studies of acyclovir treatment using a PK/PD approach is scarce [21] likely because clinical research for this purpose is extremely challenging due to ethical reasons. Alternatively, *in vitro* methods can be a feasible approach to study acyclovir PK/PD relationship for treating viral encephalitis, where future effort may be warranted.

Conventionally, whether an acyclovir dose is sufficient or not is judged based on the drug plasma concentration. However, drug concentration in brain ECF is probably more relevant for treating viral encephalitis since the viruses replicate within brain cells. We found that compared to plasma, acyclovir in the brain ECF and SAS had lower peak concentrations, but higher trough concentrations, possibly due to a slower elimination rate of acyclovir in CNS compartments. This difference yielded a higher PTA in the brain ECF and SAS than in plasma based on the C_min_>IC_50_ target, suggesting that an overdose may be derived based on PTA in plasma. When using the 50%*f*T>IC_50_ target, the current regimen appears sufficient in both plasma and brain ECF but not in SAS. These results highlight the importance to choose the right target compartment to guide dosing. Although drug concentration in ECF can be hardly measured in patients, a PBPK modeling approach can be instrumental for us to understand the drug PK in its target site so that an optimal dose decision can be made accordingly.

Toxicity is an additional aspect we considered in the acyclovir dose evaluation as acyclovir has been reported to cause neurotoxicity. Although with debate, 9-carboxymethoxymethylguanine (CMMG), an acyclovir metabolite, is believed to be responsible for the acyclovir-related neurotoxicity [22]. In this study, due to lack of data, we were not able to investigate CMMG and therefore adopted an empirical definition of acyclovir toxicity based on acyclovir plasma concentration. We found that solely increasing the dose might not maintain both efficacy and safety of acyclovir treatment, since the C_max_ in plasma following a 20 mg/kg dose would exceed the minimum toxic level of 25 mg/L [19, 23, 24]. Interestingly, in simulation we observed no drug accumulation after multiple doses with a commonly used dosing interval of 6 to 8 hours. This suggests that dosing more frequently might be safer compared to increasing the dose. Taking into account both efficacy as determined by the stricter PK target C_min_>IC_50_ for brain ECF and SAS concentrations, and toxicity, a dosing regimen of 10 mg/kg QID for HSV encephalitis and 15 mg/kg QID for VZV encephalitis appear to be both effective and safe for treating adult patients.

In this study, we used PBPK modeling combined with a fully Bayesian analysis. Such an approach allowed us to analyze sparse data and integrate existing knowledge, which was particularly suitable for studying drug treatment for CNS disease, which in our case is viral encephalitis where sampling from human is often extremely limited [17, 18, 25-27]. It is worth mentioning that the literature data we used for the modeling were not from viral encephalitis patient which may be associated with different parameter values. For this, we added a 150% to 200% uncertainty to the posterior for parameters of relevance to allow for a wider parameter space to reflect a heterogeneous population of viral encephalitis patient. Further effort might be needed in focusing on patients specifically with viral encephalitis to refine the model further for the target population. Additionally, only aggregate data were available in the identified literature, and we were not able to model the random effect to address inter-individual variability, which was necessary for individual prediction for dose individualization, e.g. for model-based therapeutic drug monitoring.

In conclusion, we have successfully developed a CNS PBPK model using Bayesian methodology for describing acyclovir CNS PK and evaluated the current and alternative acyclovir dose regimens for treating viral encephalitis. The model simulation suggested that the current standard dosing regimen of 10 mg/kg IV TID may be inadequate for treating HSV and VZV encephalitis. An alternative dose regimen of 10 mg/kg or 15 mg/kg every 6 hours may be advised considering both efficacy and toxicity. The choice of target can have an impact on the dose decision. Future effort should be made to understand the relationship between acyclovir PK and its antiviral effect to better derive dosing strategies for optimal therapeutic outcome.

## Data Availability

All data produced in the present study are available upon reasonable request to the authors

## Acknowledgement

We would like to extend our sincere gratitude to Dimitra Eleftheriou and Coen van Hasselt for their invaluable advice and insightful discussions throughout this research.

