## Supplemental file for "Revisiting acyclovir dosing for viral encephalitis using a Bayesian PBPK modeling approach"

### Supplementary document

**Figure S1. LeiCNS-PK3.0 Sensitivity Analysis Results.** The heatmap highlights the central nervous system (CNS) parameters that impact the drug pharmacokinetic (PK) profile in plasma, brain extracellular fluid (ECF), and the subarachnoid space. The colored scale represents the fold change in drug PK parameters, where a value of 1 indicates no change. The selection criteria were based on the impact of model parameter changes on the drug PK parameter area under the concentration-time curve (AUC) exceeding 15% in any model compartment ($\frac{|\mathrm{AUC}_{changed} - \mathrm{AUC}_{unchanged}|}{\mathrm{AUC}_{unchanged}} > 15\%$). Parameters include central compartment clearance (CLe), absorption rate constant (Ka), brain extracellular fluid flow (QECF), cerebral blood flow (QCBF), cerebrospinal fluid flow (QCSF), surface area of the blood-brain barrier (SABBB), surface area of the blood-cerebrospinal fluid barrier (SABCSFB), surface area of brain cells (SABC), central compartment volume (Vcen), brain extracellular fluid volume (VECF), brain cell volume (VICF), third-fourth ventricle volume (VTFV), lateral ventricle volume (VLV), cisterna magna volume (VCM), subarachnoid space volume (VSAS), lysosome volume (VLYS), brain microvessel volume (VMV), blood-brain barrier width (WBBB), blood-cerebrospinal fluid barrier width (WBCSFB), pore size of the blood-brain barrier (PZBBB), and pore size of the blood-cerebrospinal fluid barrier (PZBCSFB).

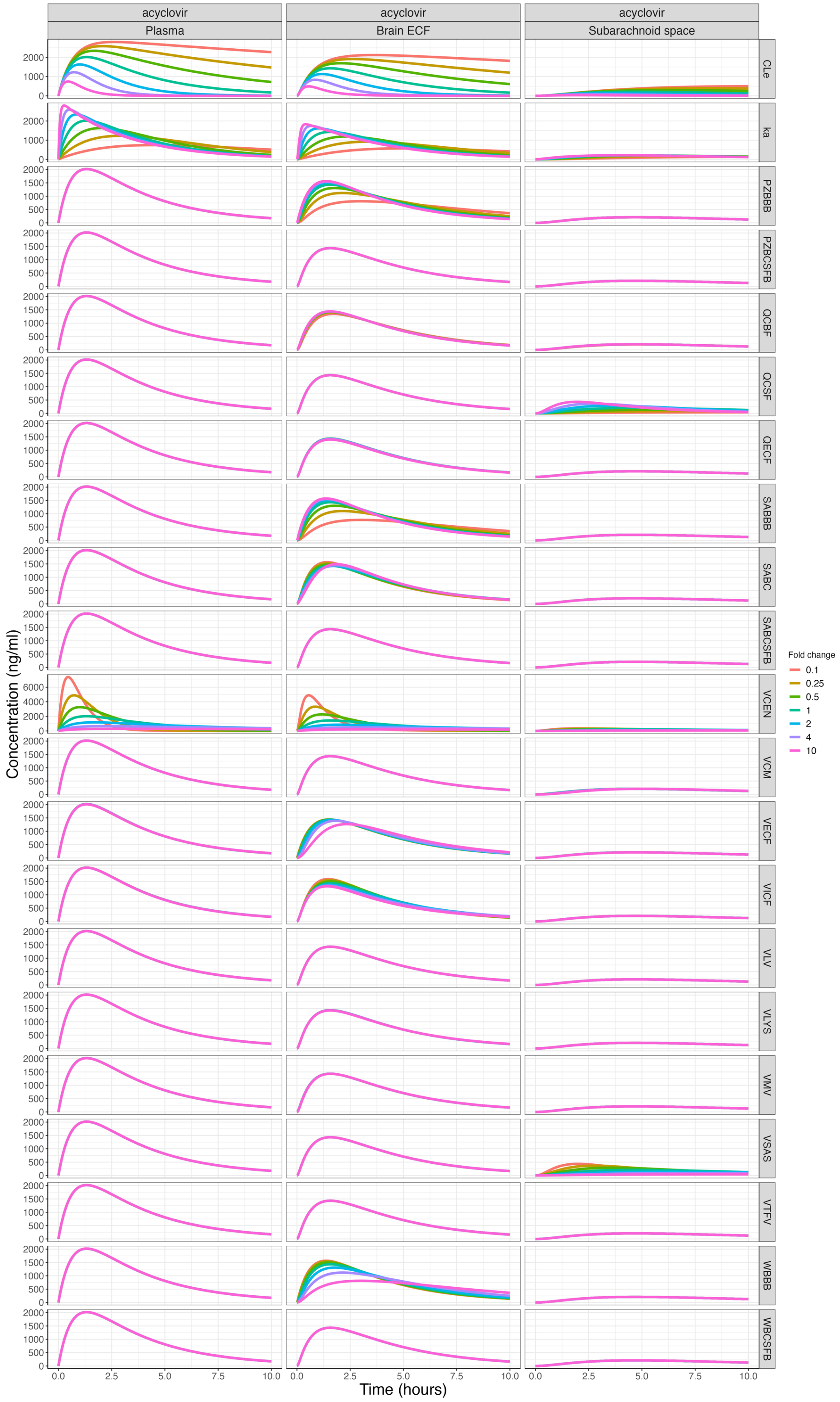
acyclovir
Plasma
acyclovir
Brain ECF
acyclovir
Subarachnoid space
CLe
ka
PZBBB
PZBCSFB
QCBF
QCSF
QECF
SABBB
SABC
SABCSFB
VCEN
VCM
VECF
VICF
VLV
VLYS
VMV
VSAS
VTFV
WBBB
WBCSFB
Concentration (ng/ml)
Time (hours)
Fold change
0.1
0.25
0.5
1
2
4
10

**Figure S2. Gelman-Rubin Statistic *R*hat and Effective Sample Size for Parameter Estimates.** (A) Gelman-Rubin statistic *R*hat values for parameter convergence, where *R*hat≤1.05 indicates satisfactory convergence. (B) Effective sample size (Neff) normalized to the total number of iterations (N) for each parameter estimate, with Neff/N > 0.5 representing sufficient sampling efficiency. Parameters include brain barrier (BBB) width (W_BBB), BBB surface area (SABBB), central compartment clearance (CLe), cerebrospinal fluid (CSF) flow rate (Q_CSF), volume of subarachnoid space (VSAS), volume of central compartment (VCEN), BBB pore size (PZ_BBB), absorption rate constant (ka) and parameters for unexplained error in plasma (Sigma_Plasma) and SAS concentrations (Sigma_SAS).

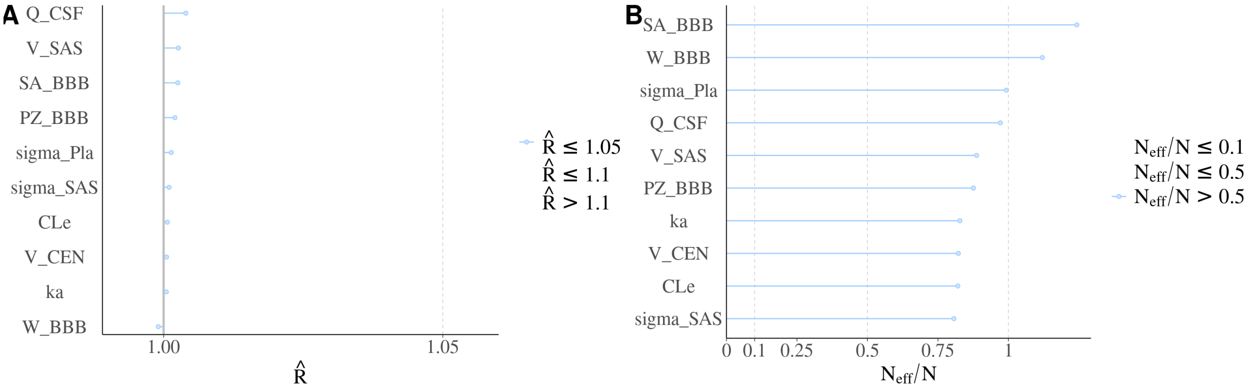

**Figure S3. Trace Plot and Posterior Distribution of Selected Model Parameters.** (A) Trace plots showing the convergence of the Markov chain Monte Carlo (MCMC) samples for each chain across selected model parameters: blood-brain barrier (BBB) width (W_BBB), BBB surface area (SA_BBB), central compartment clearance (CLe), cerebrospinal fluid (CSF) flow rate (Q_CSF), pore size of BBB (PZ_BBB), volume of subarachnoid space (V_SAS), volume of central compartment (V_CEN), and absorption rate constant (ka), along with parameters for unexplained error in plasma (sigma_Pla) and subarachnoid space (sigma_SAS) concentrations. (B) Posterior distribution of the same model parameters and parameters for unexplained error. The distributions compare the prior (yellow) and posterior (green) densities, illustrating the parameter estimates after incorporating the observed data.

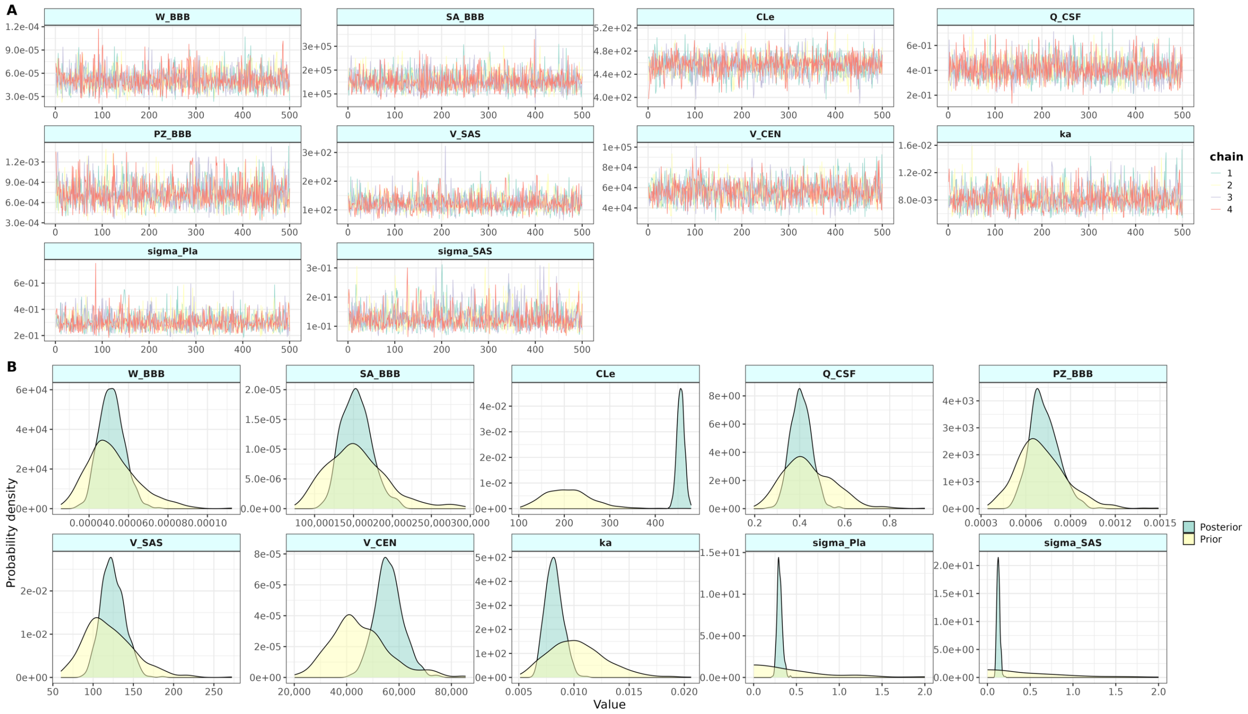

Table S1. Drug-specific and fixed systemic parameters in the model.

| Drug-specific Parameters | | Value | Reference |
|---|---|---|---|
| Molecular weight | | 225.2 | Obtained from Drugbank [28]. |
| LogP | | -0.95 | |
| pKa | | 11.98 | |
| pKb | | 3.02 | |
| Kpuu,ECF | | 0.32 | KpuuECF assume the same with rat [29], but human translation was corrected using the ratio of OAT and P-gp expression between human and rat (at least eightfold lower in human than in rat) [30]. |
| Kpuu,CSF | | 0.23 | Range from 0.19-0.27 based on $AUC_{CSF}/AUC_{serum}$ ratio [17, 18]. |
| Systemic parameters (fixed) in the model | | | |
| Plasma-related (Not used in the simulation) | | Value | Reference |
| Bioavailability of valacyclovir (F, %) | | 54.5 | Information obtained from Ref [31] |
| CNS-related | | Value | Reference |
| Volumes (mL) | Total brain ($V_{tot}$) | 1250 | Information obtained from [16]. |
| | Brain cell lysosomes ($V_{LYS}$) | 12.5 | |
| | Brain microvasculature ($V_{MV}$) | 47 | |
| | Total cerebrospinal fluid ($V_{CSF}$) | 140 | |
| | Lateral ventricles ($V_{LV}$) | 20 | |
| | 3rd & 4th ventricles ($V_{TFV}$) | 3 | |
| | Cisterna magna ($V_{CM}$) | 1 | |
| | Brain extracellular fluid ($V_{ECF}$) | 253 | |
| | Brain intracellular fluid ($V_{ICF}$) | 1000 | |
| Flow (mL/min) | Cerebral blood flow ($Q_{CBF}$) | 689 | |
| | Brain ECF bulk flow ($Q_{ECF}$) | 0.2 | |

| | | |
|---|---|---|
| Surface areas ($cm^2$) | Blood CSF barrier ($SA_{BCSFB}$) | 15000 |
| | Brain cell membrane ($SA_{BCM}$) | 2666520 |
| | Lysosomes membrane ($SA_{LYS}$) | 1980260 |
| Width (µm) | Blood CSF barrier ($W_{BCSFB}$) | 0.5 |
| Number | Total brain cells ($N_{br,cells}$) | $1.71E^{11}$ |
| Pore size (µm) | Blood CSF barrier ($PZ_{BCSFB}$) | 0.027 |
| Effective surface area (%) | BBB Transcellular transport ($SA_{BBB,T}$) | 99.8 |
| | BCSFB Transcellular transport ($SA_{BCSFB,T}$) | 99.8 |
| | BBB paracellular transport ($SA_{BBB,P}$) | 0.004 |
| | BCSFB paracellular transport ($SA_{BCSFB,P}$) | 0.016 |
| pH | Plasma ($pH_{PL}$) | 7.4 |
| | Brain microvasculature ($pH_{MV}$) | |
| | Brain extracellular fluid ($pH_{ECF}$) | 7.3 |
| | Cerebrospinal fluid ($pH_{CSF}$) | 7.3 |
| | Brain cells ($pH_{ICF}$) | 7 |
| | Brain cell lysosomes ($pH_{LYS}$) | 5 |

Table S2. Prior knowledge on the selected model parameters and unexplained errors

| Values | Mean | SD* | Ref. |
|---|---|---|---|
| Plasma parameter (Log-normal distribution) | | | |
| Central compartment clearance ($CL_e$, mL/min) | 202 | 0.25 | Information obtained from Ref [32] |
| Central compartment volume ($V_{CEN}$, mL) | 44000 | 0.25 | |
| Absorption rate constant ($K_a$, $min^{-1}$) | 0.01 | 0.25 | |
| CNS parameter (Log-normal distribution) | | | |
| Subarachnoid space volume ($V_{SAS}$, mL) | 115.83 | 0.25 | Information obtained from Ref [16]. |
| BBB pore size ($PZ_{BBB}$, μm) | 0.0007 | 0.25 | |
| BBB surface area ($SA_{BBB}$, $cm^2$) | 150000 | 0.25 | |
| BBB width ($W_{BBB}$, cm) | 0.00005 | 0.25 | |
| CSF flow rate ($Q_{CSF}$, mL/min) | 0.42 | 0.25 | |
| Values | Location | Scale | Ref. |
| Unexplained residual (Half-cauchy distribution) | | | |
| $Sigma_{Plasma}$ | 0 | 0.5 | To generate large enough tails to encompass more potential values [15]. |
| $Sigma_{SAS}$ | 0 | 0.5 | |

Table S3. Summary of sample results from parameter posterior distribution

| Variables | Mean | SD | Q5 | Q95 | $\hat{R}$ | Ess_bulk | Ess_tail |
|---|---|---|---|---|---|---|---|
| CLe (mL/min) | 456 | 17 | 428 | 482 | 1 | 1685 | 1409 |
| VCEN (mL) | 56200 | 10455 | 39991 | 73794 | 1 | 1654 | 1723 |
| VSAS (mL) | 125 | 28 | 85.3 | 175 | 1 | 1909 | 1430 |
| PZ_BBB (µm) | 0.000724 | 0.000184 | 0.000458 | 0.00107 | 1 | 1876 | 1095 |
| Ka ($min^{-1}$) | 0.0082 | 0.00155 | 0.00602 | 0.011 | 1 | 1695 | 1557 |
| SABBB ($cm^2$) | 154000 | 37809 | 101000 | 223000 | 1 | 2505 | 1380 |
| Width_BBB (cm) | 0.0000514 | 0.0000127 | 0.0000334 | 0.0000745 | 1 | 2297 | 1462 |
| QCSF (mL/min) | 0.406 | 0.0883 | 0.274 | 0.561 | 1 | 1949 | 1460 |
| Sigma_Plasma | 0.304 | 0.0607 | 0.223 | 0.416 | 1 | 2314 | 1474 |
| Sigma_SAS | 0.128 | 0.0366 | 0.0835 | 0.2 | 1 | 1958 | 1339 |
